# MUC16 mutation is associated with tumor grade, clinical features, and prognosis in glioma patients

**DOI:** 10.1101/2022.02.10.22270821

**Authors:** Valéria Pereira Ferrer

## Abstract

MUC16 is a member of the attached mucin family that encodes cancer antigen 125 (CA-125), but the association of MUC16 status with grade and subtypes of glioma patients has not yet been established. Data for MUC16 mRNA expression in 37 different cancer types were considered, and genomic data from the Cancer Genome Atlas (TCGA) from 1051 low-grade glioma (LGG) patients and 833 glioblastoma (GBM) patients were analyzed. LGG and GBM has low expression of MUC16, but it is frequently mutated in GBM. Kaplan-Meier survival analysis, glioma subtypes, methylation, and isocitrate dehydrogenase (IDH1) status were all performed. We found that mutated-MUC16 in LGG patients is associated with better prognosis considering overall survival (OS), IDH1, methylation, 1p/19q, and 10q status. Conversely, MUC16 mutation were related with worse prognosis in GBM patients upon analyzing those same parameters. Therefore, MUC16 mutations may assist in glioma diagnosis and prognosis and should be further studied in this tumor type.

## INTRODUCTION

Gliomas are tumors derived from glial cells and glial precursor cells and one of the most frequent and malignant tumors among the primary adult brain tumors (Davis 2018). Low-grade gliomas (LGG) are slow-growing tumors classified by WHO (World Health Organization) as grade I or II (diffuse gliomas) (Wang and Mehta, 2019) or III (anaplastic astrocytomas, Perry and Wesseling 2016). Grade I applies to tumors with a low proliferative rate that can potentially be cured following surgical resection. Grade II neoplasms are generally infiltrative despite having a low-level proliferative rate. Some types may progress to higher grades of malignancy. WHO grade III includes nuclear atypia and abrupt mitotic activity; these patients must receive adjuvant radiation and/or chemotherapy (Louis et al. 2007). LGG predominantly affects young adults (until 40 years-old); they are composed of various distinct tumors based on histopathology (Youland et al. 2013). More recently, the WHO reclassified LGG by combining histopathologic features with molecular markers such as isocitrate dehydrogenase (IDH) mutations and 1p/19q codeletion status. In this classification, prognosis is more strongly associated with molecular diagnostic characteristics (Louis et al. 2016). Patients with LGG generally have a favorable prognosis (Wang and Mehta 2019).

In contrast, glioblastomas (WHO grade IV) are very deadly tumors (Louis et al. 2007) and feature high rates of proliferation and invasiveness. They have high heterogeneity both inter and intra-tumor, high angiogenesis rates, and resistance to therapies (Ferrer et al. 2018; Carballo et al. 2021). The overall survival rate (OS) after diagnosis is only 12-15 months on average (Omuro and DeAngelis 2013). Thus, novel therapies to improve quality and expectation of life as well as new biomarkers to improve diagnosis and prognosis are urgently needed for this cancer type (Omuro and DeAngelis 2013; Pessoa et al. 2020).

MUC16 is a membrane-attached member of the mucin family that is expressed by some normal cells such as corneal, ovarian, and bronchial epithelial cells (Davies et al. 2007; Wang et al. 2008; Xiong et al. 2011; Felder et al. 2014). A transmembrane region of these glycoproteins’ anchors in the cell membrane, and MUC16 can be released from the cell surface by proteolytic cleavage from proteases (Hattrup and Gendler 2008). The released domain of MUC16 has a very similar molecular weight as the intact molecule (Felder et al. 2014). MUC16 is a very glycosylated mucin, and these post-translational modifications near the site of cleavage regulates release of these glycoproteins’ ectodomains (Blalock et al. 2008; Morgado and Carson 2017). Molecular cloning of MUC16 revealed that this protein has a molecular weight of 3–5 MDa (Yin and Lloyd 2001). Like other mucins, MUC16 is composed of a tandem repeat region sandwiched between the C-terminus, attached at membrane, and N-terminal domains. CA-125 is a repetitive peptide epitope of MUC16 composed of 156 amino acids (aa) (Felder et al. 2014; Morgado and Carson 2017). The MUC16 gene is present in the short arm of chromosome 19 (at 19p13.2) and is composed of 84 exons encoding for a 22,152 amino acid protein (Figure 3C). This gene is susceptible to substitutions, insertions, deletions, copy number variation (CNV) gain and losses (Figure 3D), and different methylation patterns depending on the tissue. There is no clear consensus regarding the MUC16 promoter sequence to date (Haridas et al. 2014).

MUC16/CA-125 expression has been correlated to progression and clinical status in patients with breast (Reinartz et al. 2012; Morgado et al. 2016), ovarian (Bottoni and Scatena 2015; Zhai et al. 2020), and pancreas tumors (Szwedziak et al. 2013; Thomas et al. 2021). Mutated MUC16 has already been observed and explored clinically in malignancies such as endometrium (Hu and Sun 2018), lung (Kanwal et al. 2018), melanoma (Wang et al. 2020), and gastric cancers (Yang et al. 2020). MUC16 has also gained attention as a novel target to therapy in several cancer types (Felder et al. 2014; Aithal et al. 2018).

Some members of the mucin family have been detected in gliomas (Hagel et al. 1994; Romeijn et al. 1994; Oosterkamp et al. 1997). However, to date, MUC16 mutations have not yet been explored in these tumors. Here, for the first time, we demonstrate the proof-of-principle that the MUC16 mutation has value in glioma diagnosis and prognosis. We related MUC16 mutations to molecular types and subtypes of gliomas and associated mutated MUC16 to the prognosis of these tumors. Therefore, we open an avenue and suggest that MUC16 mutations could be further studied and explored in glioma patients.

## MATERIALS AND METHODS

### Source of data analysis and mutation frequency counting

Data for MUC16 differential mRNA analysis from the 37 different tumors were obtained at the Broad Institute of Massachusetts Institute of Technology (MIT) & Harvard Portal (http://firebrowse.org/viewGene.html). Data from the TCGA database for both LGG and GBM samples were obtained from the cBio Portal (www.cbioportal.org) (Cerami et al. 2012; Gao et al. 2013).

LGG analysis selected the following data: 530 samples from TCGA Brain LGG from GDAC firehose (https://gdac.broadinstitute.org/runs/stddata 2016_01_28/data/LGG/20160128/); 514 samples from Brain LGG TCGA PanCancer Data (Bonneville et al. 2017; Ding et al. 2018; Ellrott et al. 2018; Gao et al. 2018; Hoadley et al. 2018; Liu et al. 2018; Sanchez-Vega et al. 2018; Taylor et al. 2018; Bhandari et al. 2019; Poore et al. 2020); 61 samples from a whole exome sequence study (Johnson et al. 2014); and 1102 samples from another whole exome sequencing work (Ceccarelli et al. 2016). After refinement to enrich the analysis in WHO grade III samples and compare with GBM, we selected 312 oligodendrogliomas specimens (14.0%); 212 astrocytomas (12.7%); 277 oligoastrocytomas (12.4%); 141 anaplastic astrocytomas (6.3%); and 78 anaplastic oligoastrocytomas (3.5%). We excluded 1,123 diffuse gliomas (50.4%); 13 glioblastomas (0.6%); and 1 LGG (NOS, <0.1%) from the analysis. This led to 1,051 patients and 1,090 LGG samples.

The following were selected for GBM analysis: 42 samples from a whole-genome sequence study (Zhao et al. 2019); 543 samples from a whole-exome and whole genome sequence analysis (Brennan et al. 2013); 206 samples from a targeting sequences from primary GBM in the TCGA (Network 2008); 619 TCGA samples from the GDAC firehose (https://gdac.broadinstitute.org/runs/stddata 2016_01_28/data/GBM/20160128/); and 592 GBM from the TCGA PanCancer database (https://www.cell.com/pb-assets/consortium/pancanceratlas/pancani3/index.html). Of these 2,036 samples, a refinement was performed to select samples classified as “glioblastoma” and “glioblastoma multiforme” (41.2%) excluding the samples classified as “gliomas” (58.7%). This refinement led to the 833 patients and 840 samples analyzed here.

The analysis of gene mutation and its frequency were analyzed via cBio Portal: Of the 1,090 LGG samples, 848 were profiled. Of the 840 GBM samples, 520 were profiled.

### Overall survival analysis

Overall survival data was downloaded from cBio Portal for the 848 profiled LGG samples and for the 520 profiled GBM samples. Survival data was censored until the last date that the patient was known to be alive. The samples were first segregated in a MUC16 mutated gene or MUC16 wild-type gene. Survival functions were estimated using Kaplan-Meier method and compared using a log-rank (Mantel-Cox) test in GraphPad Prism software (version 5). The median survival time was calculated as the smallest survival time for which the survivor function is equal or less than 50%. After, a derivative analysis of survival function was performed, only data with more than 50% probability of survival was used for LGG patients or less than 50% of probability of survival for GBM. A new survival curve was obtained for this analysis. Next, data from GBM patients carrying phosphatase and tensin homolog (PTEN), tumor protein (TP53), titin (TTN), epidermal growth factor receptor (EGFR), and MUC16 mutations were used to create a Kaplan-Meier survival curve and log-rank (Mantel-Cox) test. This survival graph was created in the cBio Portal.

### MUC16 mutation profile

The analysis of MUC16 mutation from 66 GBM patients was obtained in the Broad Institute of Massachusetts Institute of Technology (MIT) & Harvard Portal (http://firebrowse.org/viewGene.html) at the Tumor Portal. The types of mutation were classified in synonymous or missense mutations, in-frame or frameshift insertions/deletions and splice-site or nonsense mutations. Blue-white-red bars are log2 copy ratio distributions (**–1** to **+1**) from (Zack et al. 2013) performed at the Tumor portal site. The number of MUC16 mutations and the number of covered bases of MUC16 gene were tabulated from these GBM patients. Data from LGG patients was not disponible to compare.

### Clinical features of glioma patients

Clinical data for the IDH1 status, 1p/19q co-deletion, methylation patterns, and 10q deletion status were obtained from the cBio Portal. Clinical information from 160 GBM samples (150 patients) and from 23 LGG samples (23 patients) were also analyzed. Percentage bar graphs were obtained from these parameters after analyzing the data and using the cBio Portal.

## RESULTS

### Mutated-MUC16 is within the most frequent genes in GBM but not in the LGG patients

Analysis of 37 different types of tumors indicated that both LGG and GBM have very low MUC16 mRNA expression. LGG (194 samples) have the lowest expression with a median of -0.904 (−1.42; 0.205); GBM (33 samples) has the fourth lowest expression with a median of -0.626 (−1.04; 0.384). There was no difference between the expression of LGG+GBM specimens and normal tissue samples (Figure 1A).

**Figure 1.**
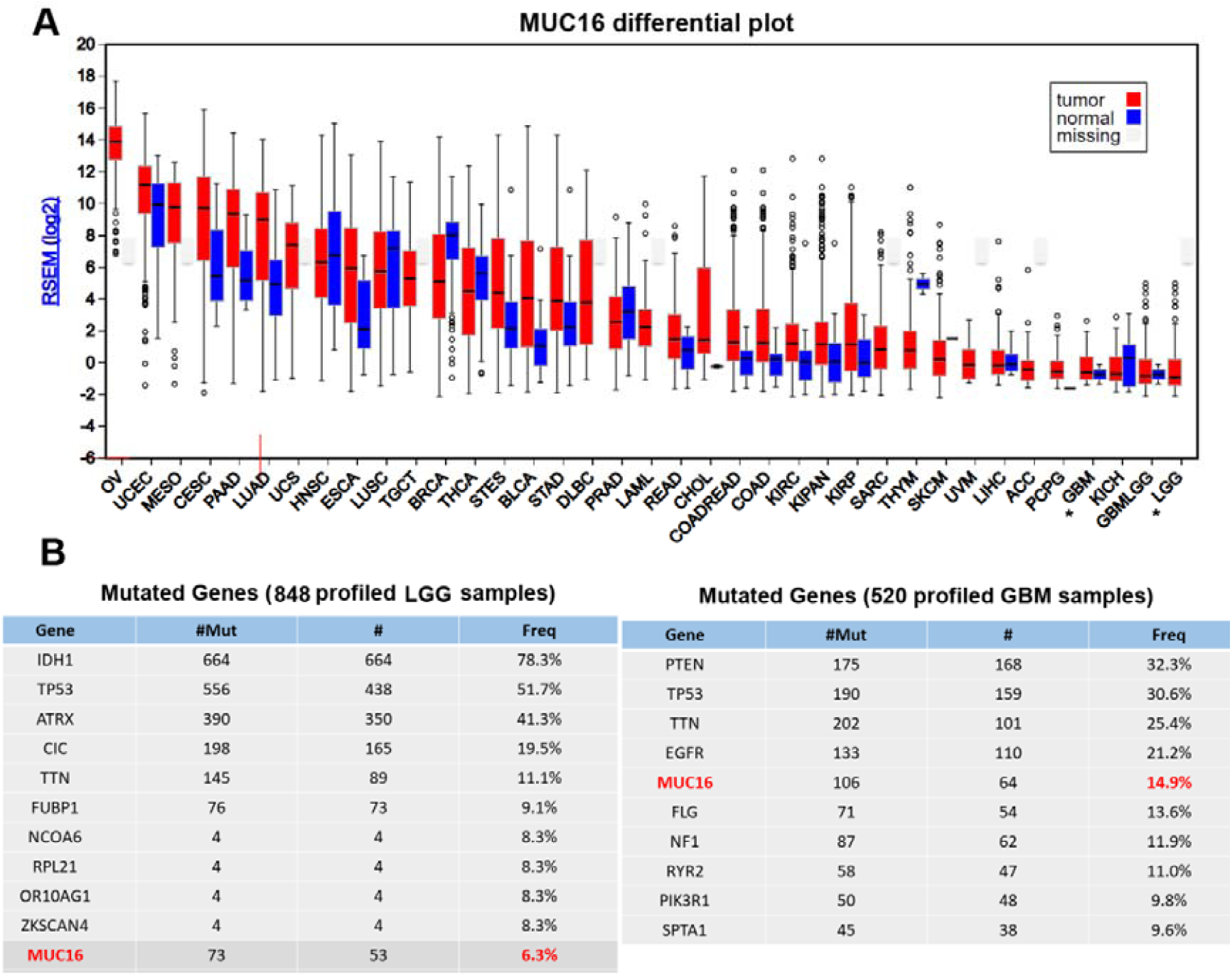
MUC16 mutation frequency but not MUC16 mRNA levels are related to glioma grade. (A) MUC16 differential plot. MUC16 mRNA differential levels in different tumor types. Ovarian and uterine cancers have the highest MUC16 mRNA levels while gliomas and kidney cancer have the lowest rates. LGG has the lowest levels. (B) List of the topmost frequent genes in LGG and GBM samples: MUC16 is within the top five most frequent gene in GBM samples, but it is not in the top ten most frequent in LGG. #Mut = total number of mutations; # = number of samples with one or more mutations; freq = frequency; OV = ovarian serous cystadenocarcinoma; UCEC = uterine corpus endometrial carcinoma; MESO = mesothelioma; CESC = cervical squamous cell carcinoma and endocervical adenocarcinoma; PAAD = pancreatic adenocarcinoma; LUAD = lung adenocarcinoma; UCS = uterine carcinosarcoma; HNSC = head-neck squamous cell carcinoma; ESCA = esophageal carcinoma; LUSC = lung squamous cell carcinoma; TGCT = testicular germ cell tumors; BRCA = breast invasive cancinoma; THCA = thyroid carcinoma; STES = esophagogastric cancer; BLCA = bladder urothelial carcinoma; STAD = stomach adenocarcinoma; DLBC = diffuse large B-cell lymphoma; PRAD = prostate adenocarcinoma; LAML = acute myeloid leukemia; READ = rectum adenocarcinoma; CHOL = cholangiocarcinoma; COADREAD = colorectal adenocarcinoma; COAD = colon adenocarcinoma; KIRC = kidney renal clear cell carcinoma; KIPAN = panCkidney cohort; KIRP = kidney renal papillary cell carcinoma; SARC = sarcoma; THYM = thymoma; SKCM = skin cutaneous melanoma; UVM = uveal Melanoma; LIHC = liver hepatocellular carcinoma; ACC = adrenocortical carcinoma; PCPG = pheochromocytoma and paraganglioma; GBM = glioblastoma; KICH = kidney chromophobe; GBMLGG = glioblastoma plus low-grade glioma; LGG = low-grade glioma. IDH1 = isocitrate dehydrogenase 1; TP53 = tumor protein 53; ATRX = alpha-thalassemia/mental retardation syndrome, nondeletion type, X-linked ; CIC = capicua transcriptional repressor; TTN = titina; FUBP1 = far upstream element binding protein 1; NCOA6 = nuclear receptor coactivator 6; RPL21 = ribosomal protein L21 ; OR10AG1 = olfactory receptor family 10 subfamily AG member 1; ZKSCAN4 = zinc finger with KRAB and SCAN domains 4; PTEN = phosphatase and tensin homolog; EGFR = epidermal growth factor receptor; FLG = profilaggrin; NF1 = neurofibromatosis type 1; RYR2 = ryanodine receptor 2; PIK3R1 = phosphoinositide-3-kinase regulatory subunit 1; SPAT1 = spermatogenesis associated 1.

In turn, when we have analyzed the frequency of MUC16 mutation in LGG patients (848 samples), the mutated-MUC16 is not one of the top ten most frequent genes (IDH1 78.3%; TP53 51.7%; ATRX 41.3%; CIC 19.5% TTN 11.1%; FUBP1 9.1%; and NCOA6, RPL21, OR10AG1, and ZKSCAN4 8.3%) with a frequency of 6.3% (Figure 1B). However, MUC16 is the fifth most frequent gene in GBM patients (520 samples) with a 14.9% frequency. More mutations are only observed with PTEN (32.3%); TP53 (30.6%); TTN (25.4%); and EGFR (21.2%) (Figure 1B).

### Mutated MUC16 patients have better OS in LGG and worse OS in GBM versus wild-type counterparts

We questioned whether MUC16 mutations are associated with the OS of glioma patients. We analyzed 45 censored patients (LGG patients) with MUC16 mutations versus 731 censored patients with wild-type MUC16. The analysis showed that the mutated-MUC16 is associated with better prognosis in LGG patients (Figure 2A). The median survival in LGG MUC16 wild-type patients was 79.93 months, and all mutated-MUC16 patients had more than a 50% probability of survival over the time observed (p = 0.0609). We then compared the same 45 censored patients containing MUC16 mutations with the 572 wild-type MUC16 patients with more than 50% of probability to survive within the time observed (Figure 2B). For this analysis, the OS was significantly higher in patients with the MUC16 mutation (p= 0.0005) versus LGG patients with wild-type MUC16 who have had a median survival of 57.88 months.

**Figure 2.**
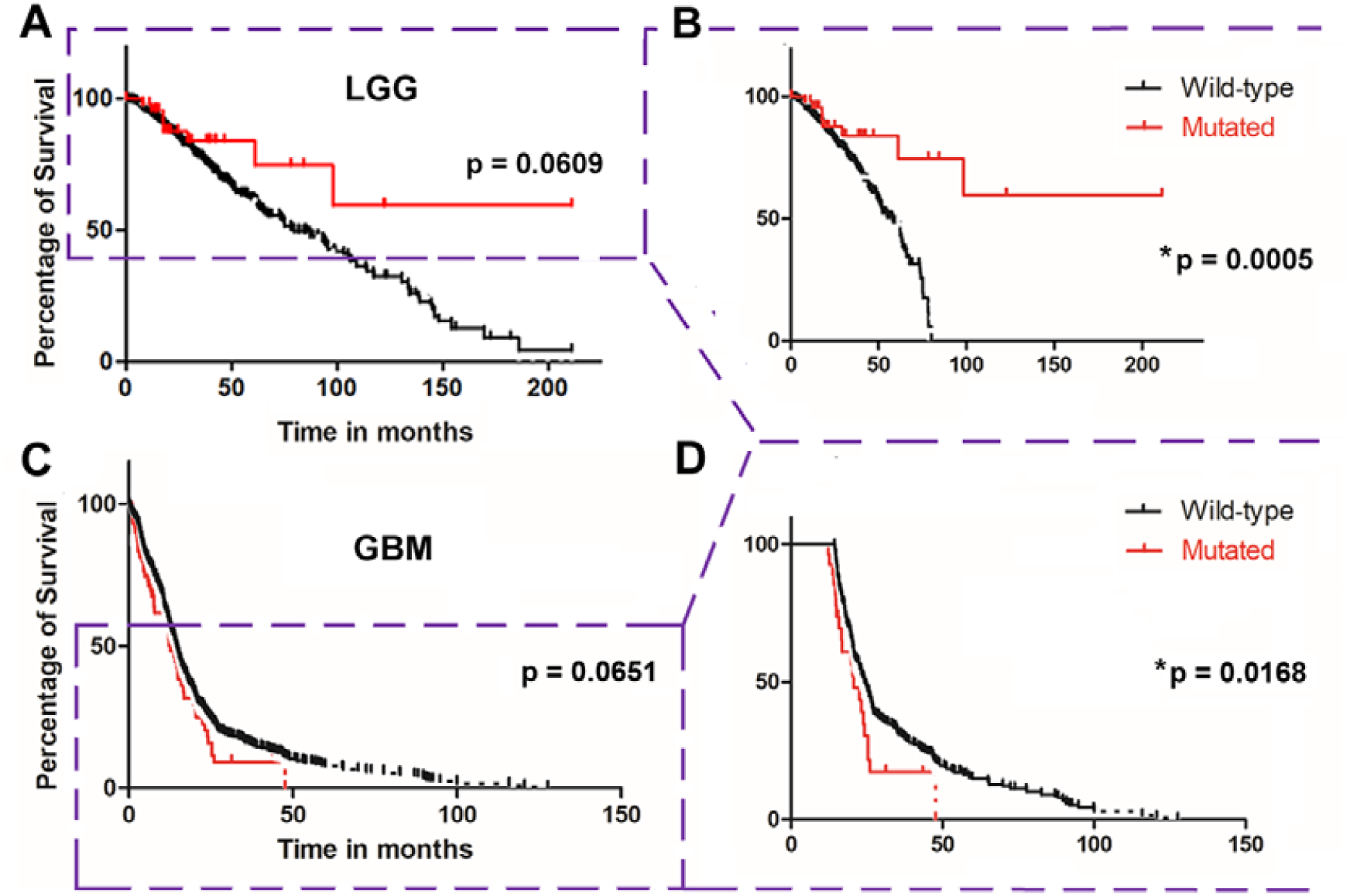
MUC16 mutations are associated with good prognosis in LGG patients and bad prognosis in GBM patients. LGG and GBM patients with mutated MUC16 and wild-type MUC16 in OS curve analysis. (A) LGG patients with a MUC16 mutation have better prognosis than their counterparts, but this is no significant (p = 0.0609). (B) In turn, among LGG patients who presented with more than 50% of probability to survive, those bearing a MUC16 mutation have better prognosis than wild-type MUC16 (*p = 0.0005). (C) Conversely, analysis of all GBM patients shows that patients bearing a MUC16 mutation have a worse prognosis than others. However, this result was not statistically significant (p = 0.0651). (D) When we have selected the GBM patients with less than 50% of chance to survive and reanalyzed the OS among them, we found that patients with mutated MUC16 have worse prognosis than wild-type patients (*p = 0.0168)

In turn, we analyzed 12 censored GBM patients with MUC16 mutations versus 133 censored patients with wild-type MUC16. Contrary to the finding in LGG patients, GBM patients with a MUC16 mutation have worse prognosis than MUC16-wild-type patients (Figure 2C): There was a median survival of 12.33 months in mutated patients versus 14.53 months in other patients (p = 0.0651). We next analyzed patients with less than 50% probability of survival, and identified six mutated MUC16 patients and 55 MUC16 wild-type patients. The OS difference was significant (p = 0.0168) with median survival times of 20.6 and 24.2 months, respectively (Figure 2D).

### Patients with a mutated MUC16 gene have a worse OS among some clinically relevant mutated genes in GBM

Among the top five most frequently mutated genes found in GBM (EGFR, MUC16, PTEN, TP53 and TTN), patients with a mutated MUC16 have the worse OS (p = 0.0074) (Figure 3A) with a median survival of 12.33 months followed by mutated EGFR (13.78 months), TTN (14.07 months), PTEN (15.02 months), and TP53 (19.82 months) (Figure 3B). Within the GBM patients with MUC16 mutation 68.2% (45/66) have missense mutation; 27.3% (18/66) have silent mutations; 3.0% (2/66) have nonsense mutation and 1.5% (1/66) have a frameshift deletion (Figure 3C). We counted the frequency of base change of missense and silent mutations in these GBM cohort. The change of guanine (G) to adenine (A) bases (G-A, 15 times) and cytosine (C) to thymine (T) bases (C-T, 13 times) were the most frequent change in the missense mutations. The change of T to C (T-C), T to G (T-G), C to A (C-A) and C to G (C-G) were present in missense mutations but not in silent mutations.

### MUC16 status correlates to clinical features of glioma patients

We next analyzed some other clinical parameters beyond OS. Data from 23 LGG patients (23 samples) and 155 GBM patients (160 samples) containing EGFR, MUC16, PTEN, TP53, and PI3K mutations were analyzed in terms of glioma type and subtype, MGMT and G-CIMP methylation status, and 1p/19q and 10q deletion. PIK3 is another clinically relevant and frequently mutated gene in GBM (Figure 2B). Figure 4A shows that ∼ 90% of LGG samples with EGFR or PTEN mutations have wild-type IDH1 while more than 60% of LGG samples with mutated PIK3 have IDH1 mutation and 1p/19q codeletion status. Among LGG mutated MUC16 patients, more than 85% of samples have an IDH1 mutation and a 1p/19q codeletion; the most of the mutated genes analyzed. All GBM samples with mutated MUC16 or with mutated PTEN have wild-type IDH1 followed by mutated EGFR (98%), PIK3 (90%), and TP53 (75%). We classified the GBM samples into recurrent or not recurrent samples (Figure 4B), and the mutated MUC16 has the second highest number of recurrency with ∼4% of samples. Mutated PIK3 is seen in ∼13% followed by PTEN (∼ 3%) and EGFR or TP53 (∼ 1%).

**Figure 3.**
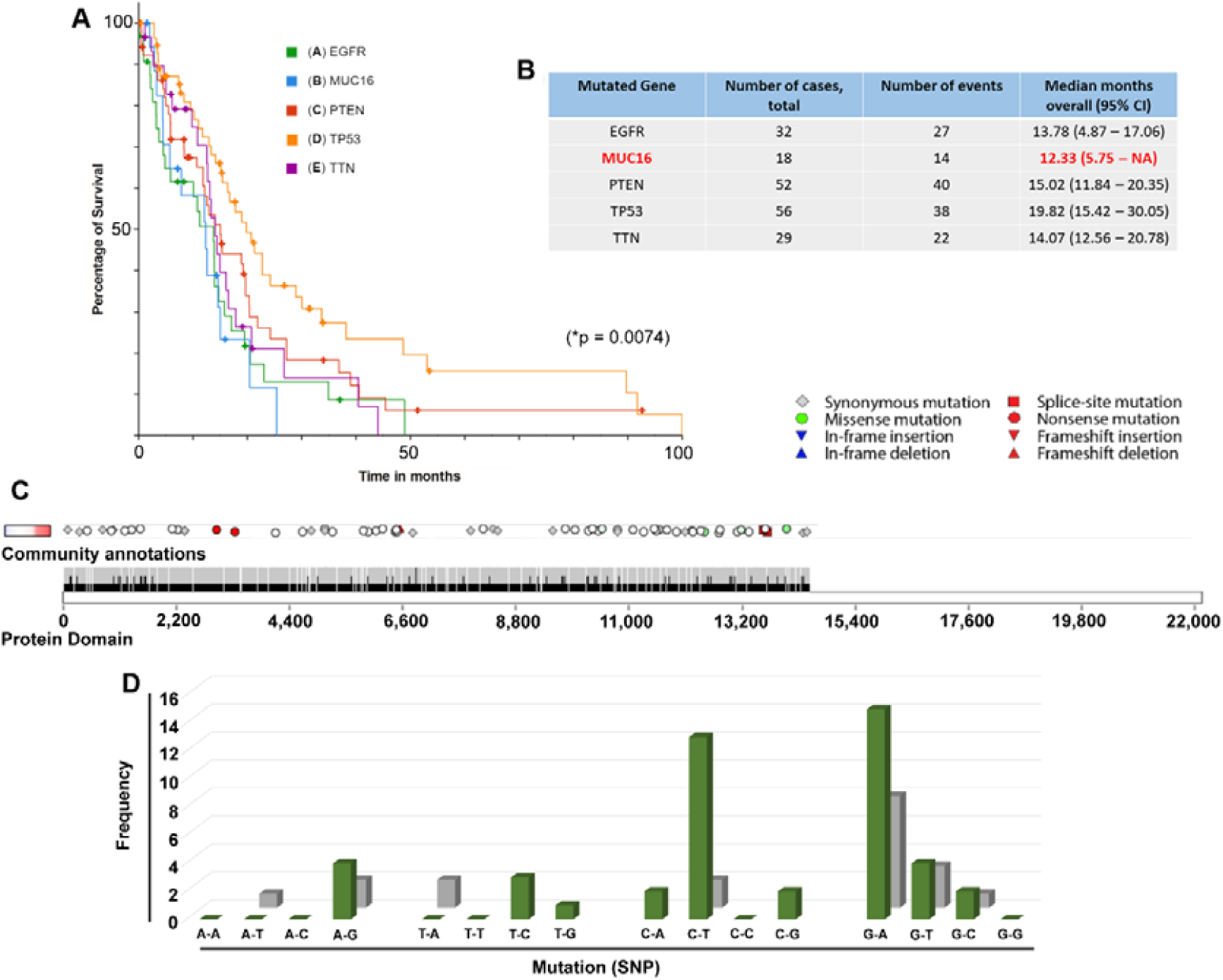
GBM patients bearing MUC16 mutation have the worse OS among clinically important mutated genes. (A) Analysis of the OS curve among GBM patients carrying EGFR, MUC16, PTEN, TP53, and TTN mutations. A worse OS is observed in patients with a MUC16 mutation followed by GBM patients with EGFR, TTN, PTEN, and TP53 mutations (*p = 0.0074). (B) Table describing the median of OS as a function of months among the five genes analyzed. (C) Graphic showing the types of MUC16 mutations in GBM cohort (N=66). Missense green saturation indicates evolutionary conservation of the mutated positions. Blue-white-red bars are log2 copy ratio distributions (**–1** to **+1**). (D) Bar graph demonstrating the frequency of base change (single nucleotide polymorphism, SNP) in missense and silence mutations (in MUC16 gene at GBM patients, N=66).

**Figure 4.**
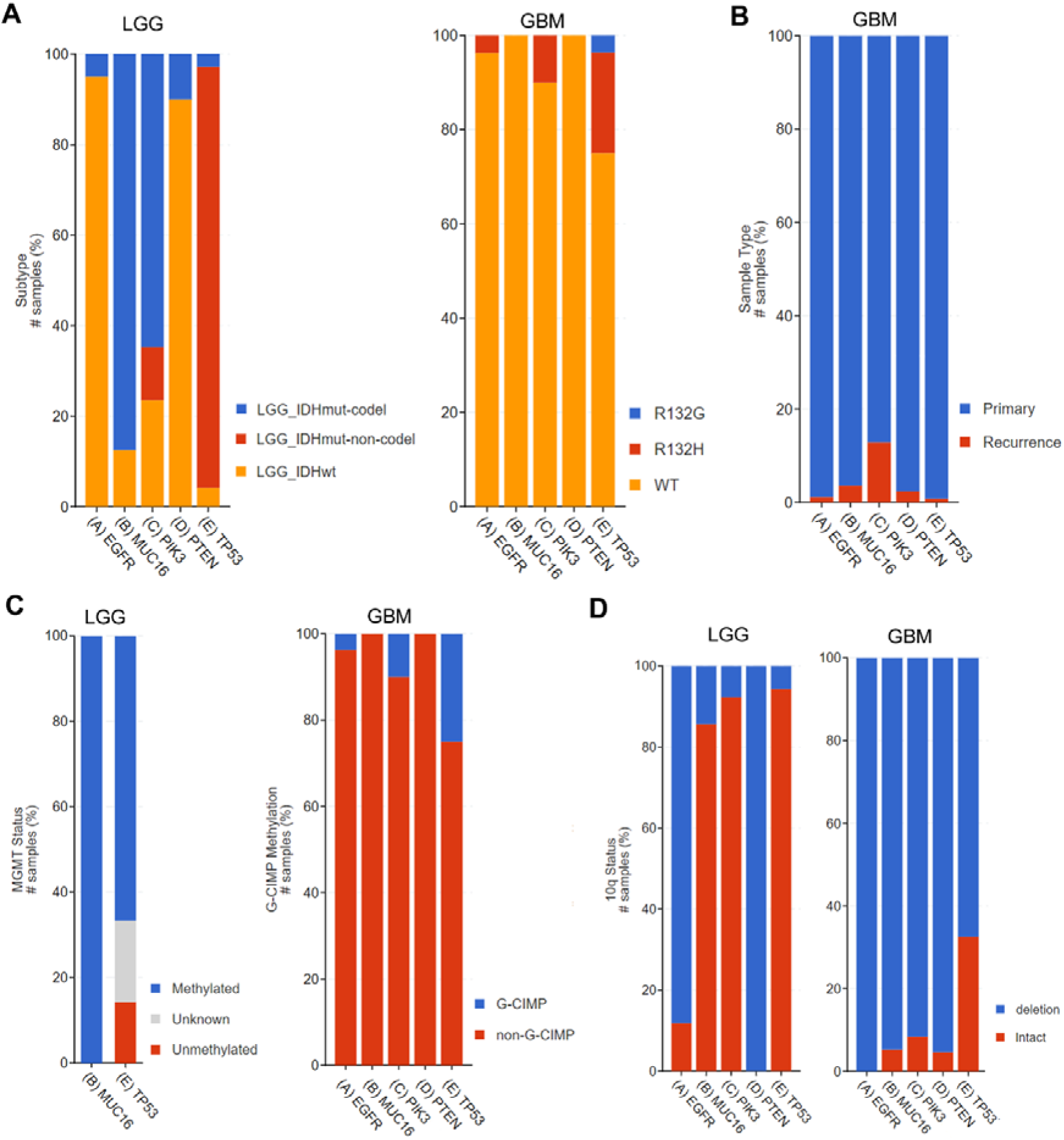
MUC16 mutations correlate to biomarkers of good outcome in LGG patients and to biomarkers of bad prognosis in GBM patients. (A) Percent of LGG and GBM patients carrying EGFR, MUC16, PIK3, PTEN, and TP53 classified by IDH1 mutation and/or 1p/19q co-deletion. Most LGG patients with MUC16 mutation have IDH1 mutated and 1p/19q co-deletion that indicates good prognosis. In contrast, all GBM patients with a MUC16 mutation have wild-type IDH1 predictive of aggressive tumors. (B) Graphic of the percentage of primary and recurrent GBM tumors classified according to the gene mutations. Most GBM patients with MUC16 mutation are primary tumors, but mutated MUC16 GBM patients have the second highest percentage of recurrence. (C) Methylation pattern of LGG and GBM patients. All LGG patients with a MUC16 mutation have a MGMT promoter methylation. This generally predicts good prognosis. All GBM patients with mutated MUC16 have non-G-CIMP methylation status that predicts worse outcomes. (D) The percentage of 10q chromosome arm deletion in LGG and GBM patients. An increased percentage of 10q deletions was observed in GBM patients versus LGG patients containing the same mutation status. 10q arm deletion has been correlated to worse prognosis in gliomas.

To classify samples based on MGMT methylation status (Figure 4C), we next obtained data from LGG-mutated MUC16 and TP53 samples. We found that 100% of mutated MUC16 samples have methylated MGMT promoter versus ∼ 65% of mutated TP53 samples. For the GBM samples we found the cytosine-phosphate-guanine (CpG) island methylator phenotype (G-CIMP) (Figure 5C); 100% of mutated MUC16 or mutated PTEN GBM samples do not have the G-CIMP phenotype. In the other GBM samples a percentage have a G-CIMP methylation status: TP53 (∼25%), PIK3 (∼10%), and EGFR (∼2%).

The chromosomal arm 10q status was studied while comparing LGG and GBM samples. For the same mutation, the percentage of chromosome 10q deletions increased significantly in GBM samples except for PTEN samples that reduced from 100% in LGG to 95% of GBM samples analyzed (Figure 4D). The mutated EGFR samples showed that the number of 10q deletions increased from 88% in LGG samples to 100% in GBM samples. The mutated MUC16 samples have a deletion that increased from 15% to 95% in LGG and GBM samples, respectively. The mutated PI3K specimens increased from 10% to 90% and from 5% to 65% in mutated TP53 samples. Therefore, patients containing MUC16 and PI3K mutations have the highest increase in 10q deletions among LGG and GBM samples.

## DISCUSSION

Gliomas account for 80% of malignant brain tumors (Schwartzbaum et al. 2006) and can be classified into LGG and high-grade gliomas (HGG), GBM. Here, LGG comprises a very diverse group of gliomas including diffuse low-grade gliomas (WHO grade II) and WHO grade III gliomas (Louis et al. 2016). Although LGG patients have better survival than GBM patients (∼7.3 years versus 18 months) (Alghamri et al. 2020), LGG can progress to GBM in some cases (Ceccarelli et al. 2016). In addition, despite the decades of research on GBM, limited progress has been made to increase the quality and expectancy of life in these patients (Shergalis et al. 2018). In this context and considering the high variability intra- and inter-evaluator on precise diagnostic based on histology alone the classification of brain tumors was revisited in 2016, and molecular criteria have been included for histopathological features (Louis et al. 2016; Chen et al. 2017). This new classification shows better compatibility between the diagnostic and the clinical evolution of patients and has opened new therapeutic possibilities. Based on a retrospective analysis of TCGA data, it has been identified important genetic mutations that assist in more precise glioma diagnosis and prognosis than histology. This includes mutations in IDH, ATRX, and TERT promoters in combination with chromosomal arm deletions of 1p and 19q among other specificities (Brat et al. 2015; Eckel-Passow et al. 2015).

The expression of MUC16 (the longest member of mucin family) has been clinically explored in several types of cancer for many years (Lakshmanan et al. 2017; Aithal et al. 2018; Fan et al. 2018, 2020; Shen et al. 2020). The classical example is ovarian cancer where it has already been used and studied for four decades (Felder et al. 2014; Coelho et al. 2018; Zhang et al. 2021). MUC16 is often one of the most frequently mutated genes in tumors (Kim et al. 2013; Wang et al. 2020; Ma et al. 2021) and is often associated with increased tumor progression (Shimizu et al. 2012; Chen et al. 2013; Das et al. 2015; Rao et al. 2015; Liang et al. 2017). However, MUC16 expression and mutation have not yet been further studied in glioma patients. Here, for the first time, we evaluated MUC16 mutation status in both LGG and GBM cohorts and correlated these with glioma grade, subtypes, and clinical outcomes.

By analyzing the expression of MUC16 in different tumors, we verified that both LGG and GBM have very low MUC16 mRNA amount; LGG has the lowest expression of all tumors analyzed. We presume that this membrane-bound mucin has not gained much attention in this type of tumor due to this low expression unlike what occurs in other types of tumors such as ovarian, endometrial, and mesothelioma cancers (Kaneko et al. 2009; Morgado et al. 2016; Liu et al. 2021) where we have observed high MUC16 mRNA expression. The analysis of other membrane-attached mucin, MUC15, has shown that its levels of mRNA and protein in glioma tissues are significantly increased versus non-cancerous brain tissues. MUC15 levels have been positively correlated with clinical stages (I, II, III vs. IV) (p <0.001) in glioma patients, and the mean value is higher in grade IV, GBM tissues (p <0.01) (Yang and Li 2014). Despite MUC16’s low mRNA expression, we have been interested in analyzing the mutation status of this gene in both LGG and GBM while searching for new diagnostic and prognostic biomarkers as well as new therapeutic targets.

Mutations in MUC16 are more common in GBM than LGG. Therefore, we hypothesized that it may be associated with progression and aggressiveness in gliomas as observed in other membrane-attached mucins, e.g., MUC1 and MUC15 (Oosterkamp et al. 1997; Yang and Li 2014). Corroborating our findings, Xiao et al. (2021) (Xiao et al. 2021) studied aging related genes as prognostic biomarkers for patients in gliomas and found that the frequency of mutations for PIK3CA, MUC16, and TTN are significantly higher in high-risk glioma cases (TTN, 26% versus 7%; MUC16, 16% versus 8%; PIK3CA, 10% versus 3%). Additionally, using multipoint sequencing for 11 patients Yang et al. (2019) (Yang et al. 2019) found that MUC16 is the most frequent gene found in GBM. The Chinese Glioma Genome Atlas GBM database shows that MUC16 mutations are higher in recurrent GBM. Here, we show that the MUC16 mutation frequency is associated with grade and malignancy in gliomas.

The MUC16 mutation status is a good prognostic biomarker in LGG patients. Similar results have been observed in other tumors. MUC16 mutations are associated with improved OS in the endometrial tumors (p = 0.0003) (Hu and Sun 2018), non-small cell lunger cancer (NSCLC) (p = 0.04), melanoma (p = 0.02) (Zhang et al. 2020), and gastric cancer cohorts (Li et al. 2018). Conversely, GBM patients with MUC16 mutations have a worse prognosis. Retrospective genome analysis agrees with our findings and demonstrated a similar result: GBM patients with a MUC16 gene mutation have worse OS outcomes than wild-type patients (p = 0.0164) (Yang et al. 2019). Here, we highlight that this result has come from GBM cohorts with less than 50% of probability to survive and that GBM patients carrying a MUC16 mutation have a worse OS among the five clinically important genes analyzed. We observed that missense mutations are the most frequent type of mutation in the MUC16 gene at GBM patients. How these MUC16 mutations culminate in the aggressive phenotype seen in GBM patients need further study.

We further analyzed the clinical aspects of glioma patients beyond OS: IDH1 status, methylation patterns, and deletion status of chromosome arms 1p/19q and 10q. Most LGG patients (65-90%) carry on IDH mutation (Sun et al. 2013; Khan et al. 2017). This type of mutation has been correlated with better prognosis in glioma cohorts who have presented better progression-free survival independent of the treatment (Wick et al. 2009; Chen et al. 2017). However, most primary GBM have wild-type IDH1, and most LGG that progresses to GBM have wild-type IDH and are aggressive tumors with worse prognosis. The presence of 1p/19q co-deletion is a strong and predictive biomarker in LGG patients (Louis et al. 2016; Chen et al. 2017).

Here, we observed that the MUC16 mutation correlates with IDH1 and 1p/19q status. Most LGG MUC16 mutation patients have IDH mutations and 1p/19q deletions, thus presenting better OS than wild-type patients. The infiltrating level of immune cells and the expression of immune checkpoint genes significantly lowers 1p/19q codeletion LGGs versus 1p/19q non-codeletion patients (Lv et al. 2021). All GBM patients carrying a MUC16 mutation have wild-type IDH with worse prognosis than non-MUC16 mutated patients. When analyzing LGG IDH1 mutated patients with high or low mutational burden, MUC16 is one of the top 10 mutated genes in the LGG IDH1 mutated patients with high mutational burden but not in the low mutational burden cohort. There were no differences in OS by analyzing MUC16 between LGG patients with high and low IDH1 mutation burden (Alghamri et al. 2020). We therefore suggest that the analysis of MUC16 mutations can be a confirmative biomarker of IDH1 and 1p/19q status, thus contributing to more precise diagnosis and prognosis in gliomas.

IDH1 mutations in gliomas have led to an increase in genome-wide methylation. This has been associated with proneural glioma subtypes with patients having a better survival outcome. A very small group of IDH-mutated gliomas lack DNA hypermethylation and have poor survival (Li et al. 2015; Chen et al. 2017; Wang et al. 2019). Thus, our results demonstrated that all LGG MUC16 mutated patients have MGMT methylation status while all MUC16 mutated GBM patients have non-G-CIMP methylation. Methylation of the MGMT promoter in the CpG-rich region is present in ∼ 40% of all GBMs and culminates in decreased expression of the MGMT protein—an enzyme that reverts the DNA alkylating effect of temozolomide (TMZ) (Hegi et al. 2005; Wick et al. 2014). Higher levels of MGMT promoter methylation, thus predicts longer survival in IDH-mutated and wild-type IDH glioma patients (Hegi et al. 2004; Fontana et al. 2016). Among people receiving radiotherapy and TMZ, MGMT promoter methylation is associated with improved median survival of 21.7 months versus 12.7 months for patients with unmethylated tumors (Stupp et al. 2009). MGMT promoter methylation is useful as a prognostic and predictive marker, but it cannot define distinct diagnostic subtypes of gliomas per se (Chen et al. 2017); thus, MUC16 status might be useful to improve diagnostic and predictive response to therapy in these patients.

Loss of heterozygosity (LOH) in 10q is common in gliomas and is associated with HGG; it is a negative prognostic marker (van Thuijl et al. 2014). The median OS for patients with or without loss of 10q is 6.6 years versus 16.7 years (p =0.009), respectively (Fontana et al. 2016). In our results, most LGG patients with MUC16, PIK3, and TP53 mutations have the 10q chromosome arm intact, which is suggestive of good prognosis and corroborates our other results. The deletion of 10q seems to be an important mechanism to progression of LGG to HGG: All GBM patients have had an increase in the deletion of this chromosome.

MGMT is located at the chromosome 10q26.3 site. Chromosome 10q LOH has a MGMT locus loss as observed in 9%, 56%, and 75% of grade II, grade III, and grade IV gliomas, respectively, thus confirming a positive correlation with tumor grade (p = 0.0002) (Sanson et al. 2002). However, there are also differences when gliomas have been classified in MGMT methylation levels according to 10q LOH. Therefore, the lack of one MGMT allele does not affect the methylation of the remaining allele (Fontana et al., 2016). We thus suggest that the measurement of frequency of the MUC16 mutation could assist in the classification of patients in LGG or GBM in combination with the 10q deletion status.

In summary, gliomas have low MUC16 mRNA expression, and GBM patients have a higher frequency of MUC16 mutations versus LGG patients. Overall, these LGG-MUC16 mutated patients have clinical features indicating good prognosis: mutated IDH1, 1p/19q chromosome arm co-deletion, high methylation patterns, low 10q deletion status, and better OS than the MUC16 wild-type patients. In turn, GBM patients carrying mutated MUC16 often have wild-type IDH1, non-G-CIMP methylation, 10q arm deletion, high rates of recurrency, and worse OS than the non-mutated patients. These clinical characteristics all predict a bad prognosis. We suggest that MUC16 gene mutation can be a new biomarker in LGG and GBM cohorts, thus assisting others classical biomarkers in finding gliomas subtypes and contributing to diagnosis, prognosis, and predicting response to therapy. We presented here a proof-of-principle conception that highlights this avenue for future work using MUC16 gene mutations as a glioma biomarker.

## Data Availability

All data produced in the present study are available upon reasonable request to the authors

## ACKNOWLEDGMENTS

The author thanks all the researchers who have contributed to the TCGA data. Acknowledgements to the researchers who built and maintained the online portals used in this work: cBio Portal; firebrowse (Broad Institute); and COSMIC online version 95. Special thanks to Dr. Micaela Morgado and Prof. Daniel Carson from Rice University (Houston, Texas) for the introduction to MUC16 biology in cancer.

## AUTHOR CONTRIBUTIONS

V.F. screened and takes responsibility for the integrity of the data and the accuracy of the data analysis, performed bioinformatic analysis and wrote the manuscript.

